# Long-COVID following mild SARS CoV-2 infection: characteristic T cell alterations and response to antihistamines

**DOI:** 10.1101/2021.06.06.21258272

**Authors:** Paul Glynne, Natasha Tahmasebi, Vanya Gant, Rajeev Gupta

## Abstract

**Background:** Long-COVID is characterised by the emergence of multiple debilitating symptoms following SARS CoV2 infection. Its aetiology is unclear, and it often follows a mild acute illness. Anecdotal reports of gradual clinical responses to histamine receptor antagonists (HRA) suggest a histamine-dependent mechanism distinct from anaphylaxis. Histamine is a paracrine regulator of T-cells: although T-cell perturbations are reported in acute COVID-19, the T-cell landscape in recovered patients and its relationship to long-COVID remains under-explored.

**Objective:** To survey T-cell populations in patients recovered from mild COVID-19, comparing those with long-COVID and asymptomatic individuals, and to analyse these data in light of symptoms and response to HRA.

**Design:** Prospective observational cohort study.

**Setting:** Single-site outpatient clinic

**Participants:** 65 (87 to 408 days post mild COVID-19). None had sought treatment for acute COVID-19. 16 recovered uneventfully (asymptomatic group), 49 presented with long-COVID (symptomatic group), of whom 25 received HRA.

**Measurements:** Structured long-COVID symptom questionnaire; quantification of T-cell subsets using a standard diagnostic assay.

**Results:** HRA significantly reduced mean symptom burden. T-cell profiles distinguished asymptomatic and long-COVID groups, but did not predict response to HRA. Long-COVID patients had reduced CD4+ and CD8+ effector memory (EM) cells and increased PD-1 expression on central memory (CM) cells. Asymptomatic controls had reduced CD8+ EM cells and increased CD28 expression on CM cells.

**Conclusion:** HRA reduce long-COVID symptoms. T-cell perturbations persist for up to 400 days following mild acute COVID-19 irrespective of long-COVID symptoms.

**Limitations:** Preliminary, single health system study.

**Primary Funding Source:** Philanthropic donations from The Dominvs Group and Sir Peter Wood

## INTRODUCTION

As of late May 2021, there have been in excess of 170 million cases of COVID-19 worldwide, with more than 3.5 million deaths (https://coronavirus.jhu.edu/map.html). Not all patients surviving the acute infection recover rapidly and uneventfully. Initial resolution of the acute illness may be followed by a combination of clinical sequelae, including but not limited to pulmonary, neurologic, dermatologic, cardiac, renal, endocrine and autoinflammatory phenomena, collectively described as long-COVID (1). Importantly, such symptoms may develop following apparent recovery from an initially mild acute illness that did not require medical intervention. It is not uncommon for at least one symptom may persist up to and beyond 7 months(2).

The majority of patients presenting with long-COVID will not have been hospitalised or sustained significant end-organ damage during their initial illness. In contrast to acute COVID-19, it predominantly affects younger patients with few comorbidities, who had relatively mild initial infectious symptoms and who did not come to medical attention until their long-COVID symptoms appeared. It is expected to place increasing burdens on healthcare systems, and by delaying the return of these often working people to normal life, it will have wider societal and economic impacts(3,4).

Long-COVID shares many features with other post-viral and idiopathic chronic fatigue syndromes(1). Persistent alterations both in the numbers of various T-Cell subtypes and their biological properties have been reported in such disorders(5,6). In acute COVID, perturbations in both B- and T-Cells are observed irrespective of disease severity(7,8). Studies of T-Cells from individuals who have recovered from COVID-19 have thus far focussed on antigen specificity, primarily to understand cellular immunity to SARS-CoV2(7,9,10). Beyond this, a detailed survey of any immunological sequelae that may persist following clearance of SARS-CoV2 and how they might relate to a cohort of long-COVID sufferers is lacking.

Whilst some patients presenting with long-COVID, particularly those who were hospitalised during their initial illness, will have clinical signs and abnormal blood or other tests, the majority do not, and there are currently no validated laboratory tests for the condition. Objective diagnostic criteria and treatment strategies are urgently required, especially as attempts to better define it remain suboptimal(4,11). Currently, several key clinicopathological questions are unanswered, specifically (i) why those individuals who go on to develop long-COVID do so; (ii) the pathological mechanisms responsible for it, and (iii) the rationale for, and efficacy of, candidate therapies.

To address these clinical imperatives, we report here the findings of a preliminary observational study undertaken in a single medical outpatient clinic to (i) describe the range of clinical symptoms in a cohort of patients presenting with long-COVID; (ii) investigate the possible benefit of HRA on long COVID symptoms; and (iii) interpret these in the light of simultaneous peripheral blood flow cytometry analysis, focusing on the numbers and phenotype of cells important to acquired antiviral immunity. All patients in the study had a mild initial infection – none had required hospitalisation for acute COVID-19, and none had received prior immunomodulatory therapy. Volunteers who had uneventfully and rapidly recovered from proven COVID infection were recruited as controls.

## METHODS

### Study setting

The Physicians’ Clinic (TPC; part of HCA healthcare UK, the study sponsor) is a private outpatient and diagnostic centre in London.

### Study design

This is a prospective observational study of patients previously diagnosed with mild COVID-19 who having initially recovered, subsequently developed persistent protean symptoms suggestive of long-COVID. Participants were recruited between November 2020 and April 2021. None had previously sought medical attention or “treatment” for acute COVID-19, none had a history of autoimmunity, and none had received immunomodulatory medications.

We recruited 49 patients with long-COVID (“symptomatic group”, symptoms >84 days following acute COVID-19 infection; physician or laboratory diagnosis) to undergo blood sampling to measure several haematological and biochemical variables, and for flow cytometry. The tests were also offered to 16 volunteer clinician colleagues, all of whom had either PCR or serological evidence of COVID-19 and who had recovered uneventfully (“asymptomatic group”). The age and gender distributions of the two groups were similar A symptom questionnaire was designed with a binary symptom grid to initially record the presence (score 1) or absence (score 0) of the following long-COVID symptom categories: fatigue, constitutional upset (sweats, fever, arthralgia, myalgia), breathlessness, post-exertional malaise (PEM), chest pain, neurological (headaches, neurosensory, brain fog), neuropsychiatric (anxiety, insomnia), dysautonomia (postural tachycardia), ear, nose and throat symptoms, gastrointestinal disturbance (food intolerance, diarrhoea, bloating), and dermatological manifestations (rashes, flushing, urticaria), to give a maximum possible symptom score of 11. The questionnaires were collated by one of us (NT) who was not involved in the management of the patients, and blinded to their clinical details.

The study protocol, patient consent form, information leaflets and questionnaire were submitted to the host Institutions’ IRB and approved. Patients and volunteers who consented to participate were subsequently fully informed of the analysis results, which were discussed face to face.

Initially patients were offered supportive care only. However, as the study progressed, all patients were offered empiric treatment trials with a combination of H1 (Loratidine 10 mg twice daily or Fexofenadine 180 mg twice daily) and H2 (Famotidine 40 mg once daily or Nizatidine 300 mg once daily) receptor antagonists (HRA) for a minimum of 4 weeks as part of their on-going care. Of the 49 long COVID study participants, 25 patients consented to try to HRA. Between 4 -16 weeks after starting treatment, both HRA-treated and untreated patients were asked to grade their symptoms as now absent, identical, better, or worse. In this analysis ‘absent’ or ‘better’ scored 0, and ‘identical’ or worse, 1.

### Flow cytometry and additional laboratory measurements

This was performed on peripheral blood collected in EDTA. The Beckman Coulter TQ-prep whole blood lysis system (Beckman Coulter Life Sciences, High Wycombe, UK) was used to prepare cells for flow cytometry. Antibody staining was with the Duraclone IM T lyophylised antibody panel (B53328, Beckman Coulter) as described previously(12). This is in routine diagnostic use in our clinical practice, and normal ranges had been previously established (in 2018) in healthy adults with normal automated blood counts for accreditation purposes. Analysis of primary flow cytometry data was in Kaluza C (Beckman Coulter) and was undertaken by one of us (RG), who was blinded to clinical information collected and collated by others. The gating strategy and phenotypes analysed are shown in Supplemental Figures 1 and 2. An automated full blood count was performed on every specimen in parallel to flow cytometry. T-Cell populations were quantified as a percentage of total cellularity, and absolute numbers calculated from the corresponding total white cell count. Single cell antigen density was recorded as median fluorescence intensity (MFI).

### Statistical analysis

Comparison of clinical data from the asymptomatic and symptomatic groups was by Mann-Whitney “t” test. Comparison of response to HRA was by Wilcoxon matched pairs signed rank test. All flow cytometry fluorescence data and numbers of T-Cell sub-populations were assumed to lie in non-Gaussian distributions, and analysis of variance was by Kruskall-Wallis H-Test. Dunn’s Multiple Comparison Test was then used to estimate statistical significance. All statistical analysis was in Prism 9.1 (GraphPad Software, LLC).

## RESULTS

### Patient characteristics

49 patients with a diagnosis of long-COVID were recruited. 25 had either PCR or serological evidence for COVID-19. The remainder had suffered their acute illnesses at the start of the pandemic, when PCR testing was not widely available in the UK, and had not subsequently returned positive serology tests. Only 1 had been vaccinated at the time of enrolment into the study. We recruited 16 individuals known to have had acute COVID infection but who had recovered rapidly and uneventfully to serve as asymptomatic controls. The majority in this group are healthcare professionals; all had had either positive PCR tests or serology, and 14 had received at least one vaccination dose (Pfizer) at the time of enrolment.

Participants’ baseline clinical features are shown in Table 1. Almost all long-COVID patients were polysymptomatic (95.8%) as described by others (13), with an average of 4.58/11 typical symptoms (range 1 – 10). The average symptom duration was 268.9 days (range 87-402) at the time of referral and participation in the study. Long-COVID patients were relatively young (mean 43; range 25 – 65 years) and showed a female preponderance (29/49; 60.4%). 17/49 long-COVID patients had a history of atopy, which is in keeping with reports that atopy is predictive of mild acute disease(14)

**Table 1.** Baseline characteristics of the study participants.

| Clinical characteristics | Long-COVID<br>(Symptomatic, n= 49) | post-COVID controls<br>(Asymptomatic, n = 16) |
| --- | --- | --- |
| Age range (median) | 25 – 65 (43) | 25-72 (34.5) |
| Female (%) | 30 (61.2%) | 8 (50%) |
| Ethnicity |  |  |
| White | 45 (91.8%) | 12 (75%) |
| Asian | 2 (4.1%) | 2 (12.5%) |
| Black | 1 (2%) | 1 (6.3%) |
| Mixed | 1 (2%) | 1 (6.3%) |
| Comorbidities<br>1 – cancer, 1 – controlled hypertension | 2 (4.1%) | 0 |
| Allergy or atopy | 16 (32.7%) | 1 (5.8%) |
| Mean days from acute COVID to study testing | 271.8 days | 321.6 days |
| Long COVID symptoms |  |  |
| Fatigue | 36 (73.5%) | N/A |
| Constitutional | 31 (63.3%) | N/A |
| Breathlessness | 19 (38.8%) | N/A |
| Post-exertional malaise | 35 (71.4%) | N/A |
| Chest pain (non-cardiac) | 18 (36.7%) | N/A |
| Neurological/neurosensory | 39 (79.6%) | N/A |
| Neuropsychiatric | 29 (59.2%) | N/A |
| Dysautonomia | 14 (28.6%) | N/A |
| Ear, nose and throat | 22 (44.9%) | N/A |
| Gastrointestinal | 20 (40.8%) | N/A |
| Dermatological | 23 (46.9%) | N/A |

### Routine Blood Tests

These were undertaken at first presentation to our clinic, and included full blood count, erythrocyte sedimentation rate, C reactive protein, D-dimer, renal and liver function tests. The results of the were within the normal range in almost every case. 2/49 patients had mildly elevated CRP (NR = <5mg/L) and 4/49 had a mildly elevated ESR (NR = <15mm/Hr). Circulating total and differential leucocyte numbers were normal in all patients; one patient had an incidental borderline normocytic anemia.

### Treatment with HRA

At first presentation, symptoms were categorised using the symptom grid. 25 patients (16 female, 9 male, mean age 44 years) were treated empirically with HRA, and 24 (14 female, 10 male, mean age 41) either declined HRA or were not offered them because they were first seen before HRA treatment became part of our practice. All patients were offered standard supportive care and advice to control symptoms (NICE guideline NG188: https://www.nice.org.uk/guidance/ng188/RCGP/SIGNguidelines).

The symptom profiles in the treated and untreated cohorts were similar (Supplemental Figure 3). HRA treatment reduced average symptom burden by 59.7% (Figure 1a). The mean time to response was 29.6 days (median 26 days; range 6 – 89 days). 5 patients (20%) reported complete resolution of all symptoms, 13 (52%) experienced some improvement, 6 reported no change, and one deteriorated, (developing PEM and insomnia shortly after starting Loratidine and Famotidine). Patients reported improvements in all symptoms except dysautonomia (Supplemental Figure 4). There was no correlation between SARS-CoV2 antibody status and response to HRA. Of the 17 long-COVID patients with a history of atopy, 11 received HRA, and of these 8 reported a clinical improvement.

**Figure 1.**
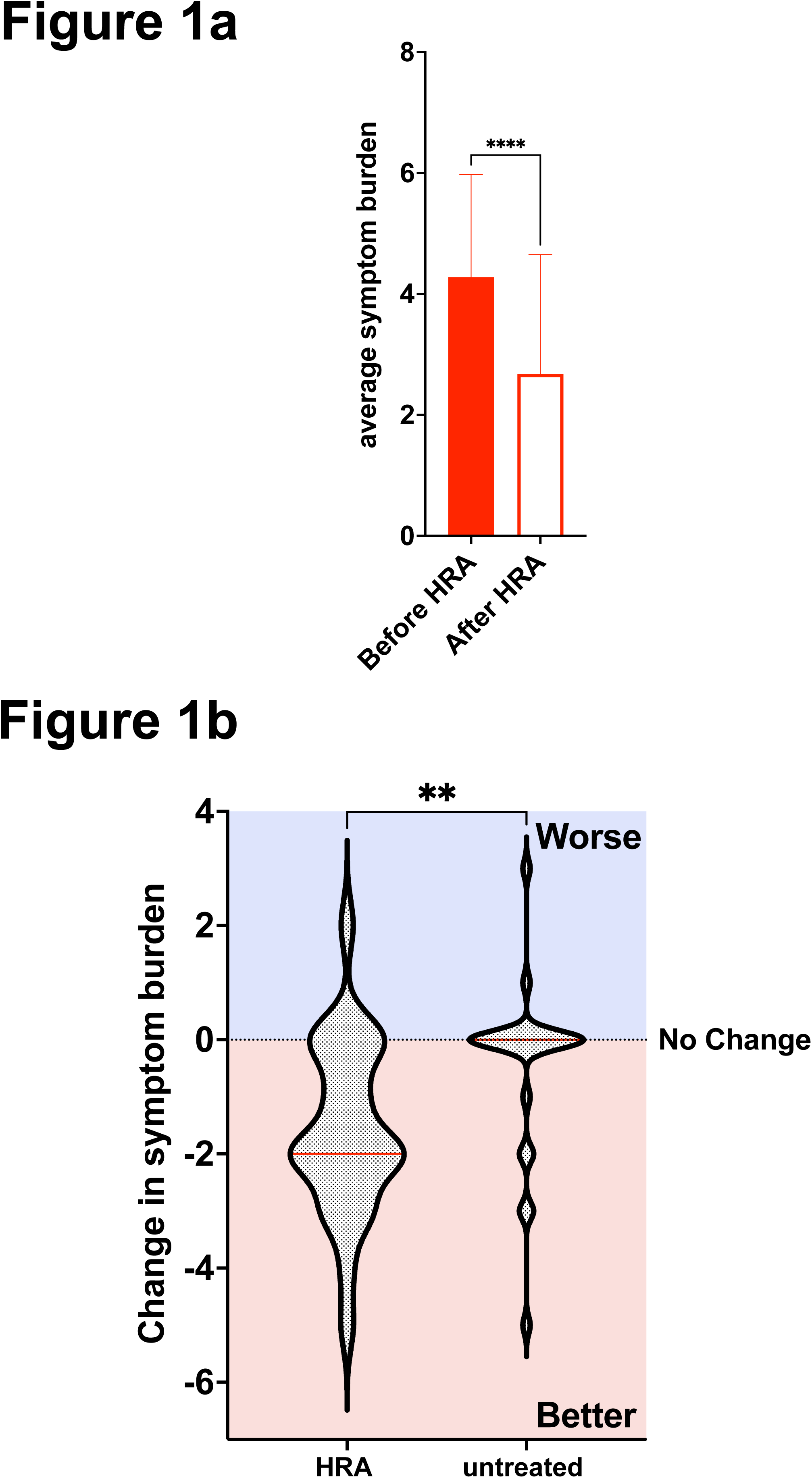
Response to HRA in symptomatic patients. **(a)** Mean symptom burden (± SD) in symptomatic patients before and after HRA treatment (n=25). Before treatment symptom range 1-8, mean 4.28/11 ± 1.7 after treatment range 0-6, mean 2.68/11 ± 1.9. p<0.0001, Wilcoxon matched-pairs signed rank test. **(b)** Change in symptom burden in symptomatic patients who did (n=25) and did not (n=24) receive HRA. Red line denotes median. HRA 59.75% reduction, untreated 10.6%. HRA: pre-treatment symptom burden range 1 – 8, mean 4.28 ± 1.7; post treatment symptom burden range 0 – 6, mean 2.68 ± 1.9. Untreated: pre-treatment symptom burden range 1 – 10, mean 4.91 ± 2.6; symptom burden range at follow up 0 – 8, mean 4.39 ± 2.6. (p=0.0052, Mann-Whitney test).

The 24 patients who did not receive HRA were also reassessed between 28 and 119 days after their initial blood tests (median 56 days). 24% reported some spontaneous improvement in their symptoms. One experienced a spontaneous full recovery, and 5 reported a reduction in some but not all symptoms. However, the majority (16, 67%) reported no change, and 2 (8%) developed new additional symptoms (Figure 1b). There was no correlation between SARS-CoV2 antibody status and spontaneous resolution of symptoms in this group.

### T-Cell compartments

Flow cytometry was performed once as part of the initial blood tests. Despite the time that had elapsed from the acute COVID-19 illness (see Table 1, Long-COVID symptomatic 87 - 408 days, asymptomatic controls 100 - 404 days), we observed marked perturbations in the numbers of circulating effector memory (EM) T-Cells.

25/49 symptomatic and 3/16 asymptomatic participants had CD4+ EM counts that were below the lower limit of the normal range (Figure 2a), and receiver operating characteristic (ROC) analysis confirmed that the CD4+ EM count could distinguish the two groups (Figure 2b). 43/49 long-COVID patients and 14/16 of the asymptomatic group had reduced CD8+ EM counts, which were below the median of the normal range (Figure 3a). Although the mean count was lower in the long-COVID than in the asymptomatic group, the CD8+ EM count did not distinguish the two cohorts in a ROC analysis (Figure 3b). All other T-Cell compartments, including CD4+ and CD8+ central memory (CM) cells, were within normal limits.

**Figure 2.**
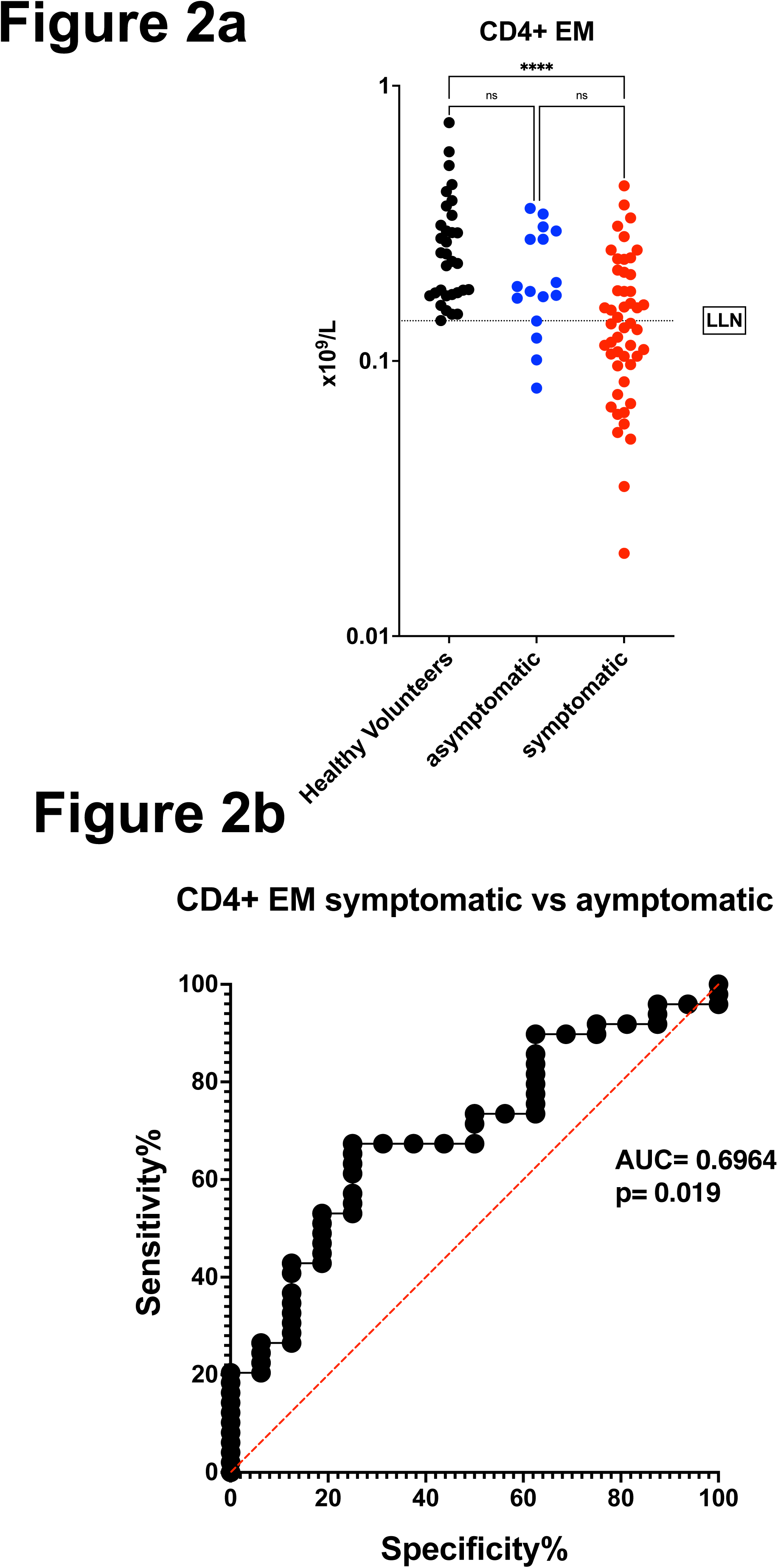
Reduced CD4+ EM T cells distinguish long-COVID patients from the asymptomatic fully recovered group. **(a)** Circulating CD4+ Effector Memory T cells (×10^9^/L) in healthy volunteers (black), asymptomatic recovered (blue) and symptomatic long-COVID (red) participants. Dashed line, LLN, lower limit of normal. Healthy volunteers: mean 0.276×10^9^/L, range 0.140×10^9^/L – 0.735×10^9^/L. Asymptomatic: mean 0.211 ×10^9^/L, range 0.079×10^9^/L – 0.359 ×10^9^/L. Symptomatic: mean 0.154 ×10^9^/L, range 0.020 ×10^9^/L – 0.433 ×10^9^/L. p (healthy volunteers *vs* symptomatic) = <0.0001, p (asymptomatic *vs* symptomatic) = 0.073 (Kruskall-Wallis test). **(b)** Receiver operating characteristic curve of CD4+ EM T cell number in symptomatic (long-COVID) and asymptomatic participants. Red dashed-line, random classifier. AUC, area under curve (c-statistic).

**Figure 3.**
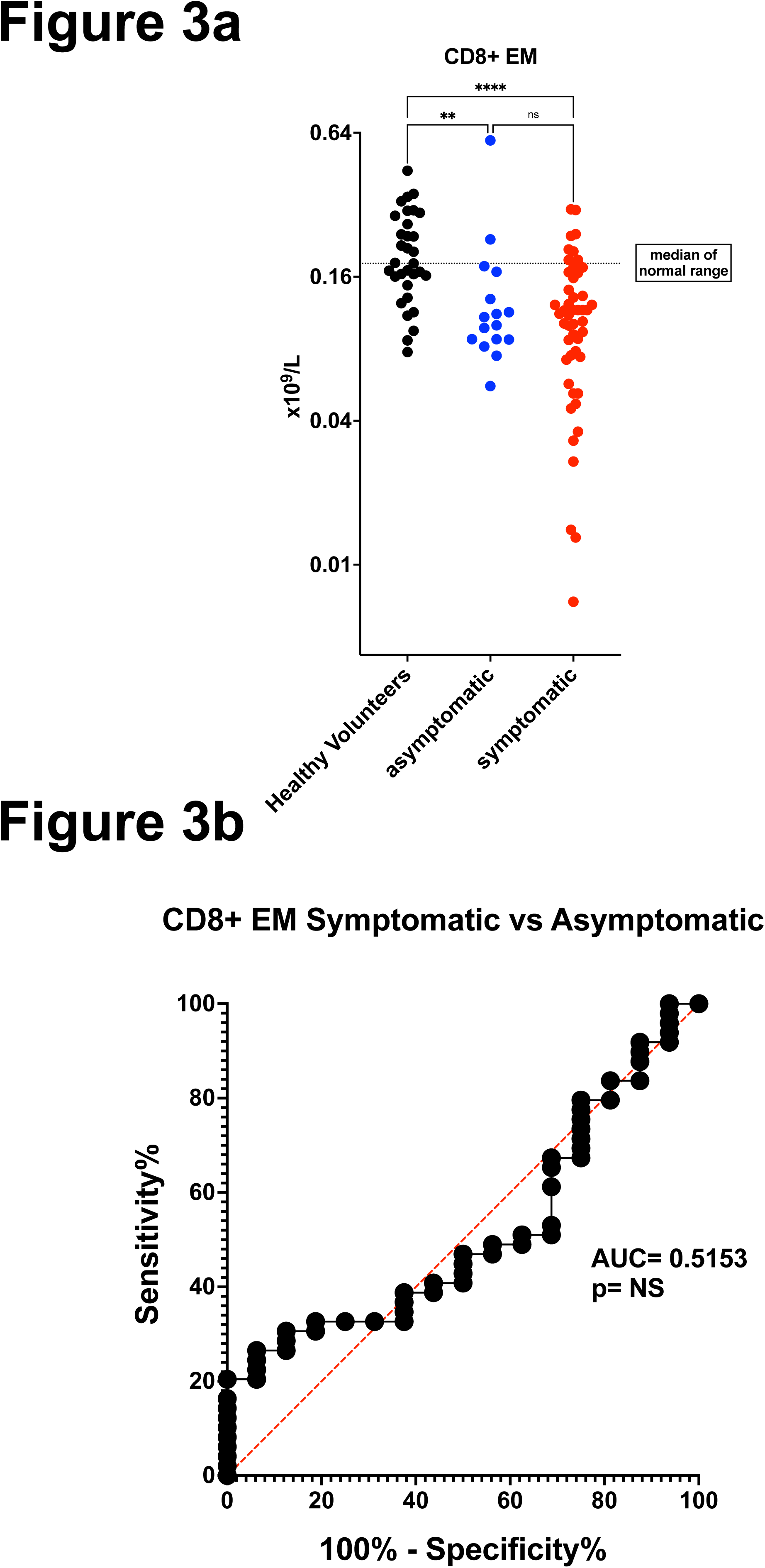
CD8+ EM T-Cell counts remain low after COVID-19 irrespective of symptoms. **(a)** Circulating CD8+ Effector Memory T cells (×10^9^/L) in healthy volunteers (black), asymptomatic (blue) and symptomatic (red) participants. Dashed line, median of normal range. Healthy volunteers: mean 0.209×10^9^/L, range 0.077×10^9^/L – 0.444×10^9^/L. Asymptomatic: mean 0.144 ×10^9^/L, range 0.056×10^9^/L – 0.594 ×10^9^/L. Symptomatic: mean 0.117 ×10^9^/L, range 0.007 ×10^9^/L – 0.306 ×10^9^/L. p (healthy volunteers *vs* symptomatic) = <0.0001, p (healthy volunteers *vs* asymptomatic) = 0.045 (Kruskall-Wallis test). **(b)** Receiver operating characteristic curve of CD8+ EM T cell number in symptomatic (long-COVID) and asymptomatic participants. Red dashed-line, random classifier. AUC, area under curve (c-statistic).

Our antibody panel allowed us to compare the expression levels (antigen densities) of proteins important for regulating T-Cell function(15). We observed that the antigen density of PD-1 (Programmed cell death protein 1, CD279) was significantly increased in both CD4+ and CD8+ CM cells in all participants, although this was more marked in symptomatic long-COVID patients (Figure 4a). Intriguingly, CD28 expression was significantly increased in CD4+ CM cells in the asymptomatic group, but not the long-COVID group (Figure 4b). Expression of both proteins was similar in all other T-Cell compartments, and expression of CD57 did not vary significantly.

**Figure 4.**
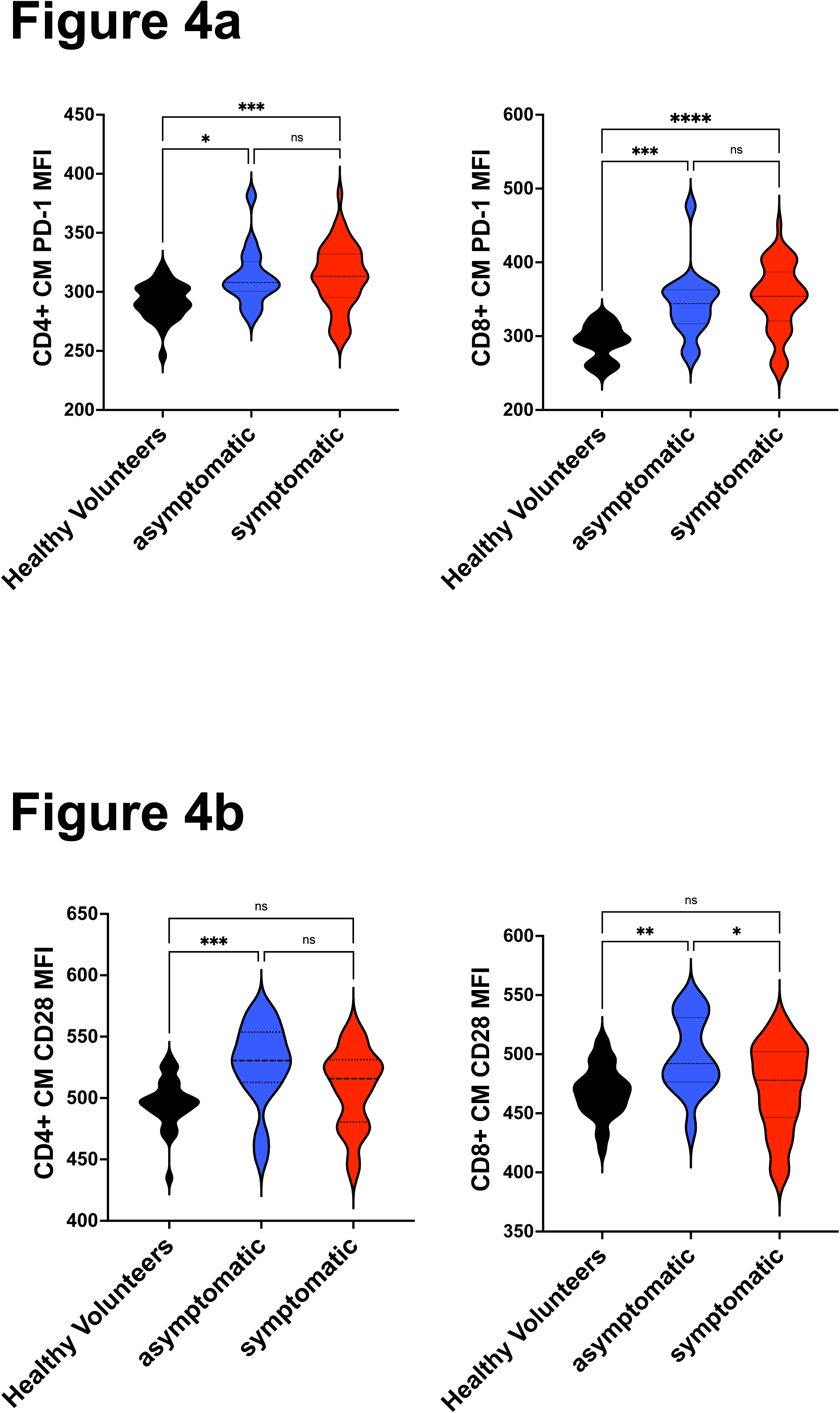
CM T cell PD1 and CD28 levels distinguish long-COVID patients from the asymptomatic fully recovered group. **(a)** PD1 antigen density expressed as median fluorescence intensity (MFI, arbitrary units) in CD4+ (left) and CD8+ CM T cells. **CD4+ CM**: Healthy volunteers, mean 292AU, range 246-321AU. Asymptomatic, mean 313AU, range 279-381AU. Symptomatic, mean 313AU, range 259-383AU. p (healthy volunteers *vs* symptomatic) = 0.0002, p (healthy volunteers *vs* asymptomatic) = 0.024 (Kruskall-Wallis test). **CD8+ CM**: Healthy volunteers, mean 292AU, range 249-329AU. Asymptomatic, mean 344AU, range 274-477AU. Symptomatic, mean 352AU, range 256-451AU. p (healthy volunteers *vs* symptomatic) = *<*0.0001, p (healthy volunteers *vs* asymptomatic) = 0.0006 (Kruskall-Wallis test). **(b)** CD28 antigen density in CD4+ (left) and CD8+ CM T cells. **CD4+ CM**: Healthy volunteers, mean 496AU, range 435-532AU. Asymptomatic: mean 528AU, range 453-572AU. Symptomatic: mean 508AU, 437-563AU. p (healthy volunteers *vs* symptomatic) = 0.068, p (healthy volunteers *vs* asymptomatic) = 0.0006 (Kruskall-Wallis test). **CD8+ CM**: Healthy volunteers: mean 469AU, range 420-512AU. Asymptomatic: mean 499AU, range 438-547AU. Symptomatic: mean 472AU, range 395-532AU. p (symptomatic *vs* symptomatic) = 0.0402, p (healthy volunteers *vs* asymptomatic) = 0.0087 (Kruskall-Wallis test).

Although both the numbers of circulating EM T-Cells, and combined PD1 and CD28 antigen density in CM cells distinguished asymptomatic from symptomatic participants (Figure 2b, Supplemental Figure 5), neither predicted responsiveness to HRA.

## DISCUSSION

The majority of patients with long-COVID will have had a mild or asymptomatic initial infection, and many are eventually diagnosed several months later. In this preliminary study, we report the clinical and immunological features of 49 such patients, none of whom had received prior medical treatment for COVID-19, comparing them with an age-matched cohort of patients who made full and uneventful recoveries from COVID-19. We have classified their symptoms and documented their clinical response to readily available low-cost medications (combined HRA), and quantified their peripheral blood T-Cells using an assay that is in routine use in a local diagnostic laboratory.

In keeping with other early series, we saw a preponderance of younger females with few comorbidities, although 35% had a past history of atopy. The majority were polysymptomatic and presented up to 400 days after their initial infection. Our key observations are that (i) While in the untreated group some 25% of patients’ symptoms improved over 4-17 weeks without specific pharmacological intervention, in the HRA-treated group significantly more (72%) reported a reduction in the number of symptoms. (ii) Average CD8+ EM T-Cell counts remained low for up to 400 days following COVID-19 irrespective of symptoms (total automated lymphocyte counts were normal). (iii) Symptomatic long-COVID was in addition associated with significantly lower CD4+ EM T-Cell counts. (iv) Levels of PD-1 were increased on CD4+ and CD8+ CM T-Cells in all participants (more marked in long-COVID), although CD4+ and CD8+ CM T-Cell counts were normal. (v) CD28 levels were higher on the CD4+ CM cells of the asymptomatic recovered group.

The slow clinical response of long-COVID to HRA does not support a classical anti-anaphylactic mechanism of action. Both CD4+ and CD8+ T-Cells express H1 and H2 histamine receptors, each of which modulates lymphocyte function *via* distinct intracellular pathways(16). Furthermore, Famotidine may also contribute to T-Cell desensitisation by stabilising H2 receptor conformation through beta-Arrestin (17,18). Accordingly, we postulate that HRA may reduce long-COVID symptoms by blocking the histamine-dependent paracrine regulation of T-Cell function. HRA improved all symptoms except dysautonomia (postural tachycardia syndrome, POTS), suggesting that this particular feature of long-COVID is driven by another mechanism. In support of this notion, COVID is increasingly recognised as triggering loss of tolerance to autoantigens (19,20), and we note that dysautonomia has been associated with both antecedent viral infections and autoantibodies to adrenergic and cholinergic receptors(19).

To our knowledge this is the first report of a rapid turnaround, routine laboratory test detecting persisting abnormalities in the circulating T-Cell landscape many months after mild COVID infection, irrespective of whether or not patients had fully recovered. Our data suggest a late, chronic phase of the T-Cell response to SARS-CoV2, perhaps linked to the earlier multi-specific and cytotoxic CM and EM responses seen in the acute infection(21,22). We found significantly lower numbers of circulating CD8+ EM cells in both our asymptomatic and long-COVID cohorts. Interestingly, in patients with acute COVID infection, dominant CD8+ T-Cell responses correlate with milder disease, suggesting a protective role in the suppression of pathogenic inflammatory responses (7,9). In contrast, our observations were made several months after the acute illness had resolved, and suggest that reduced numbers of CD8+ EM cells at this stage are linked to the recovery from COVID-19 infection itself, rather than to the development of long-COVID.

We also observed several changes restricted to patients with long COVID, including lower numbers of CD4+ EM cells, and an increased expression density of PD1 on both CD4+ and CD8+ CM cells. PD-1 acts as a co-inhibitory molecule for several CD4+ and CD8+ T-Cell functions, contributes to immunological memory(23) and is associated with T-Cell apoptosis and exhaustion in the context of chronic viral infection(24). Our additional finding of increased expression density of CD28, a co-stimulatory molecule present on both CD4+ and CD8+ T-Cells essential for signal transduction and triggering, was limited to those individuals who had made an uneventful recovery from COVID infection, and this phenomenon may represent a longer term, “healthy” and proportionate immune response to SARS-CoV2 infection.

Finally, our detection of changes in T-Cell landscape so late after infection suggest the possibility that SARS-CoV2 could persist for longer than originally assumed. Only further work will determine whether this might be associated with preliminary *in vitro* observations of the potential for reverse transcription of SARS-CoV2 RNA and genomic integration in human cells(25).

Rather than being hypothesis-driven, this was a “real life” study prompted by the clear, emerging clinical imperative presented by long-COVID, as well as suggestions that HRA may be effective in reducing symptoms, which in turn may relate to measurable, objective abnormalities in circulating T-Cell landscape. As a preliminary observational report from a single-centre, it has several limitations. However, our intriguing observations merit refinement in future prospective studies exploring the clinical signal of HRA response together with a more detailed investigation of the mechanisms underlying long term abnormalities in the T-Cell landscape in long-COVID. Finally, and with further development in a larger cohort, flow cytometric analysis may provide a rapid and straightforward diagnostic test for long COVID itself.

## Data Availability

Any data that DO NOT contain patient identifying information are available on request

## LEGENDS TO SUPPLEMENTAL FIGURES

**Supplemental Figure 1** Gating strategy used to identify T cell compartments

**Supplemental Figure 2** T cell compartments that were quantified, and their defining surface phenotypes.

**Supplemental Figure 3** Symptom profile of long-COVID patients who received HRA, and those who did not (untreated group)

**Supplemental Figure 4** Reduction in each symptom type following treatment with HRA. Neurology and neurosensory (69% reduction/resolution), dermatology (66.6% reduction/resolution), chest pain (53.8% reduction/resolution), neuropsychiatry (42.8% reduction/resolution), gastrointestinal (37.5% reduction/resolution), fatigue (31.2% reduction/resolution), constitutional (30% reduction/resolution), ENT (20% reduction/resolution), breathlessness (20% reduction/resolution) and post-exertional malaise (8.3% reduction/resolution),

**Supplemental Figure 5** Scatter plots showing CD28 and PD1 antigen density in CD4+ (left) and CD8+ EM cells following mild COVID-19. Symptomatic (Long-COVID) and asymptomatic recovered group as indicated. ULN, upper limit of normal. LLN, lower limit of normal.

