## Supplemental Figure 1 for "Long-COVID following mild SARS CoV-2 infection: characteristic T cell alterations and response to antihistamines"

### Slide 1
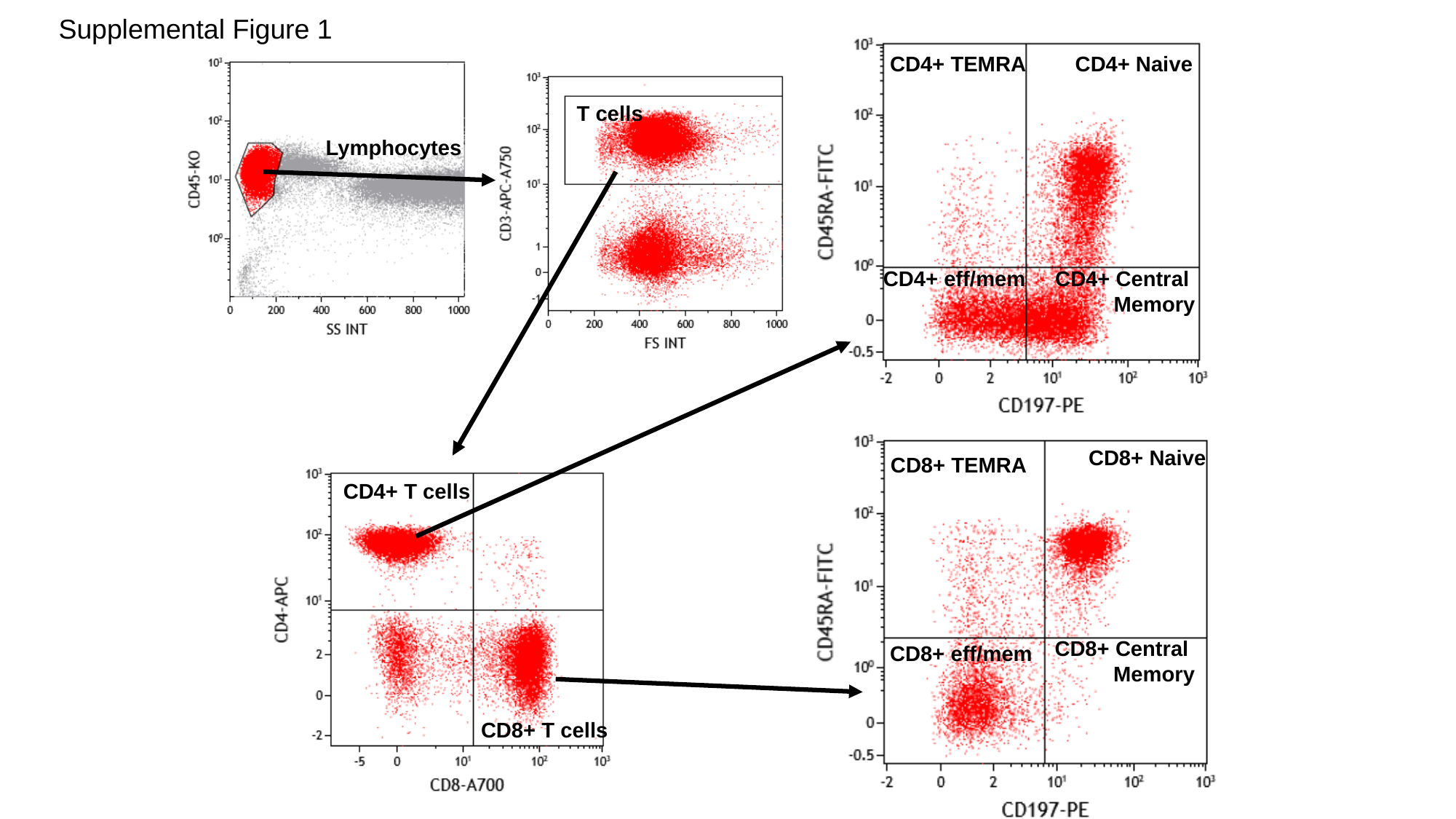

Supplemental Figure 1
CD4+ TEMRA
CD4+ Naive
CD4+ eff/mem
CD4+ Central
Memory
T cells
Lymphocytes
CD8+ Naive
CD8+ TEMRA
CD8+ Central
Memory
CD8+ eff/mem
CD4+ T cells
CD8+ T cells
