## Supplemental Figure 2 for "Long-COVID following mild SARS CoV-2 infection: characteristic T cell alterations and response to antihistamines"

### Slide 1
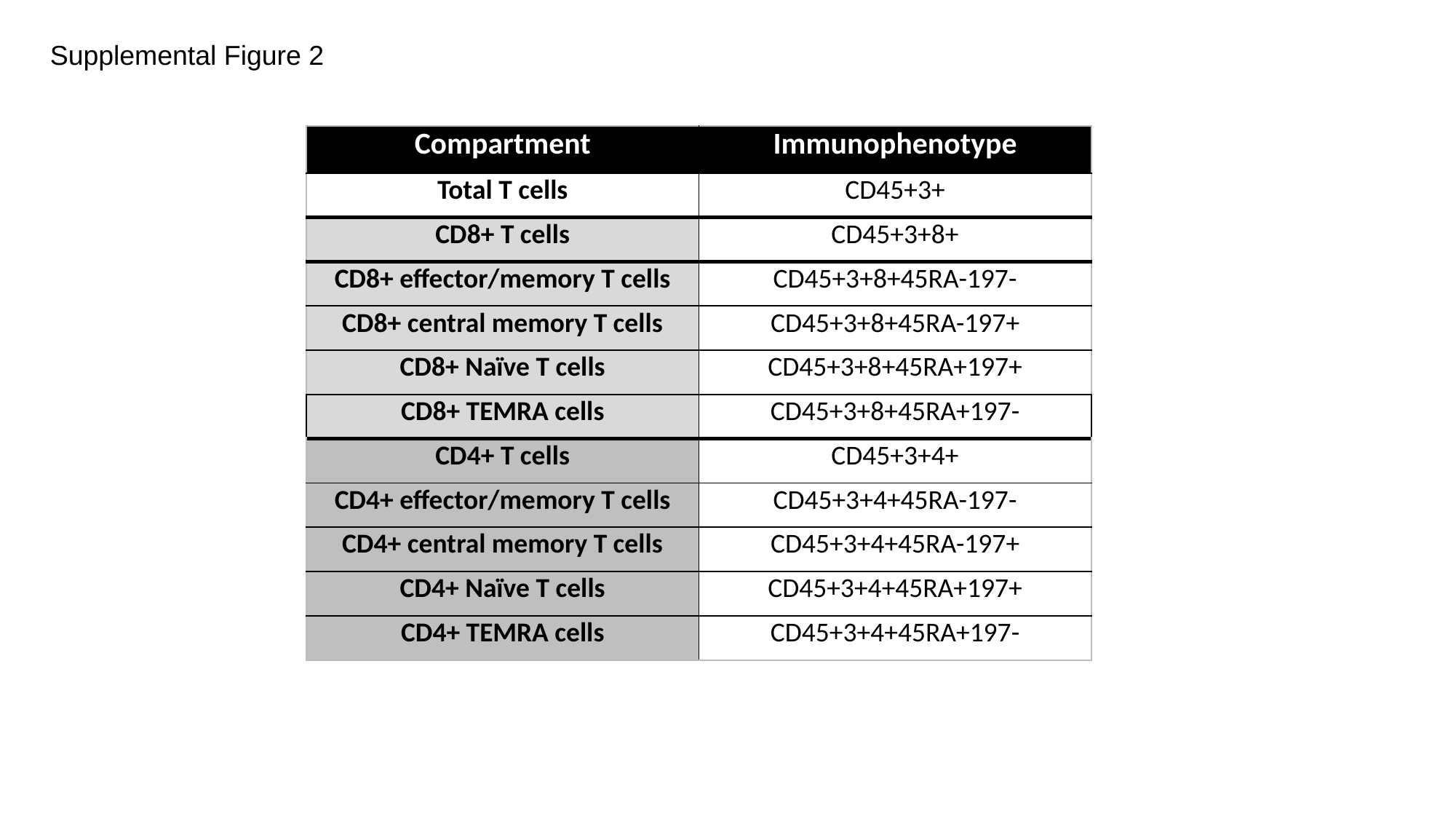

Supplemental Figure 2
| Compartment | Immunophenotype |
| --- | --- |
| Total T cells | CD45+3+ |
| CD8+ T cells | CD45+3+8+ |
| CD8+ effector/memory T cells | CD45+3+8+45RA-197- |
| CD8+ central memory T cells | CD45+3+8+45RA-197+ |
| CD8+ Naïve T cells | CD45+3+8+45RA+197+ |
| CD8+ TEMRA cells | CD45+3+8+45RA+197- |
| CD4+ T cells | CD45+3+4+ |
| CD4+ effector/memory T cells | CD45+3+4+45RA-197- |
| CD4+ central memory T cells | CD45+3+4+45RA-197+ |
| CD4+ Naïve T cells | CD45+3+4+45RA+197+ |
| CD4+ TEMRA cells | CD45+3+4+45RA+197- |
