## Supplemental Figure 3 for "Long-COVID following mild SARS CoV-2 infection: characteristic T cell alterations and response to antihistamines"

### Slide 1
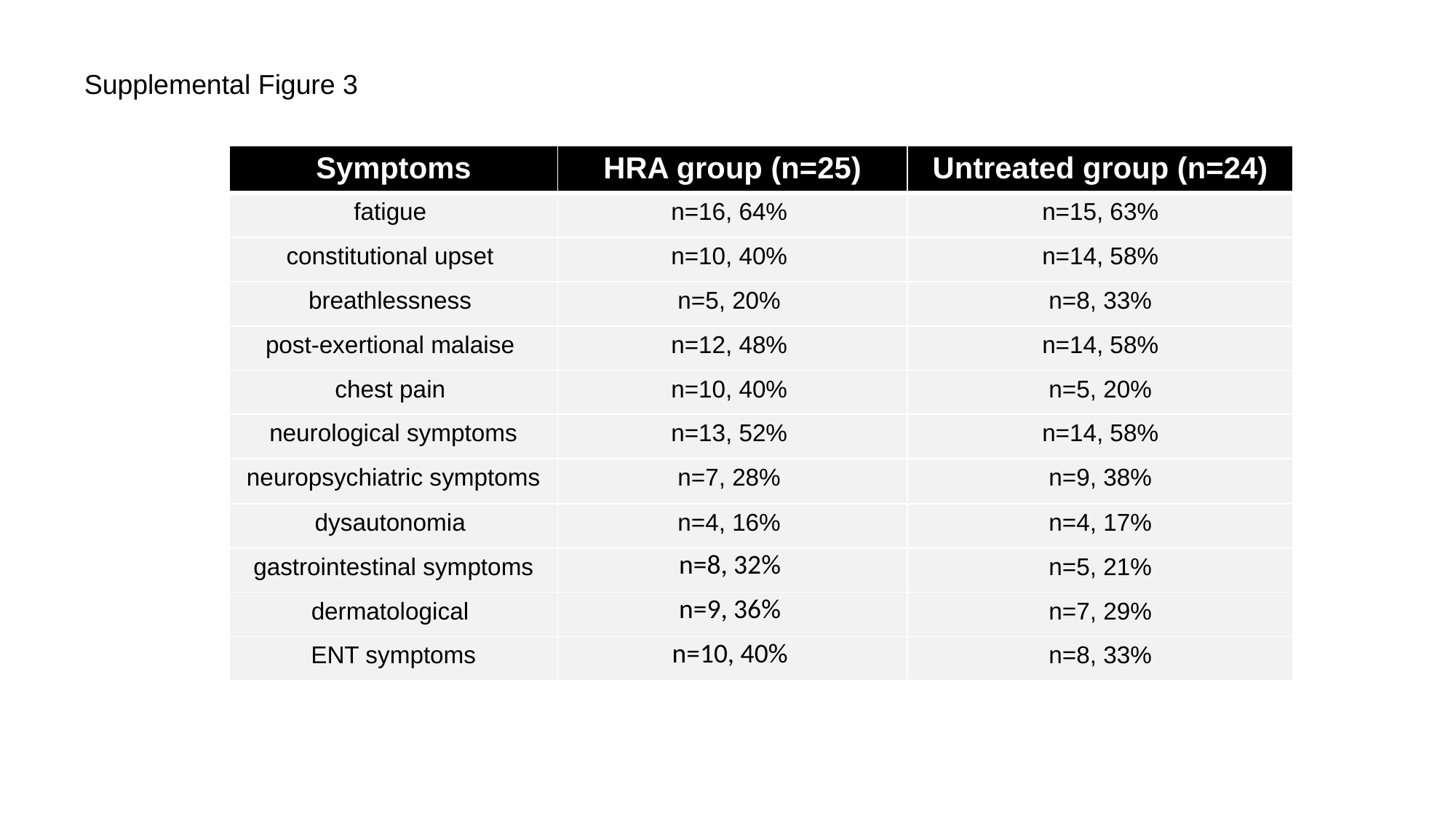

Supplemental Figure 3
| Symptoms | HRA group (n=25) | Untreated group (n=24) |
| --- | --- | --- |
| fatigue | n=16, 64% | n=15, 63% |
| constitutional upset | n=10, 40% | n=14, 58% |
| breathlessness | n=5, 20% | n=8, 33% |
| post-exertional malaise | n=12, 48% | n=14, 58% |
| chest pain | n=10, 40% | n=5, 20% |
| neurological symptoms | n=13, 52% | n=14, 58% |
| neuropsychiatric symptoms | n=7, 28% | n=9, 38% |
| dysautonomia | n=4, 16% | n=4, 17% |
| gastrointestinal symptoms | n=8, 32% | n=5, 21% |
| dermatological | n=9, 36% | n=7, 29% |
| ENT symptoms | n=10, 40% | n=8, 33% |
