## Supplemental Figure 4 for "Long-COVID following mild SARS CoV-2 infection: characteristic T cell alterations and response to antihistamines"

### Slide 1
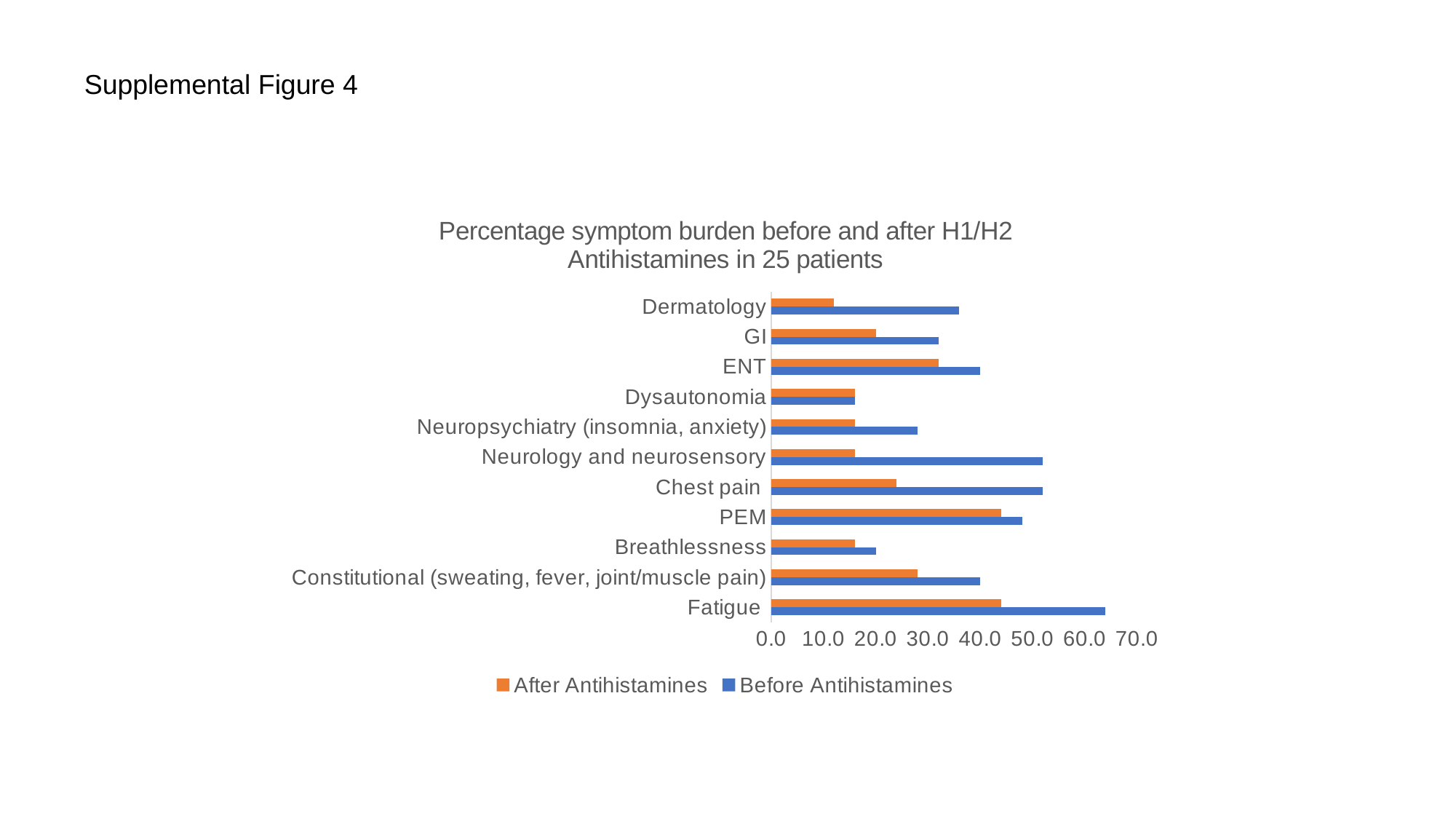

Supplemental Figure 4
#### Chart: Percentage symptom burden before and after H1/H2 Antihistamines in 25 patients
| Category | Before Antihistamines | After Antihistamines |
|---|---|---|
| Fatigue | 64.0 | 44.0 |
| Constitutional (sweating, fever, joint/muscle pain) | 40.0 | 28.0 |
| Breathlessness | 20.0 | 16.0 |
| PEM | 48.0 | 44.0 |
| Chest pain | 52.0 | 24.0 |
| Neurology and neurosensory | 52.0 | 16.0 |
| Neuropsychiatry (insomnia, anxiety) | 28.0 | 16.0 |
| Dysautonomia | 16.0 | 16.0 |
| ENT | 40.0 | 32.0 |
| GI | 32.0 | 20.0 |
| Dermatology | 36.0 | 12.0 |
