## Supplementary figures and images for "Long-COVID following mild SARS CoV-2 infection: characteristic T cell alterations and response to antihistamines"

### Supplemental Figure 5

#### Slide 1
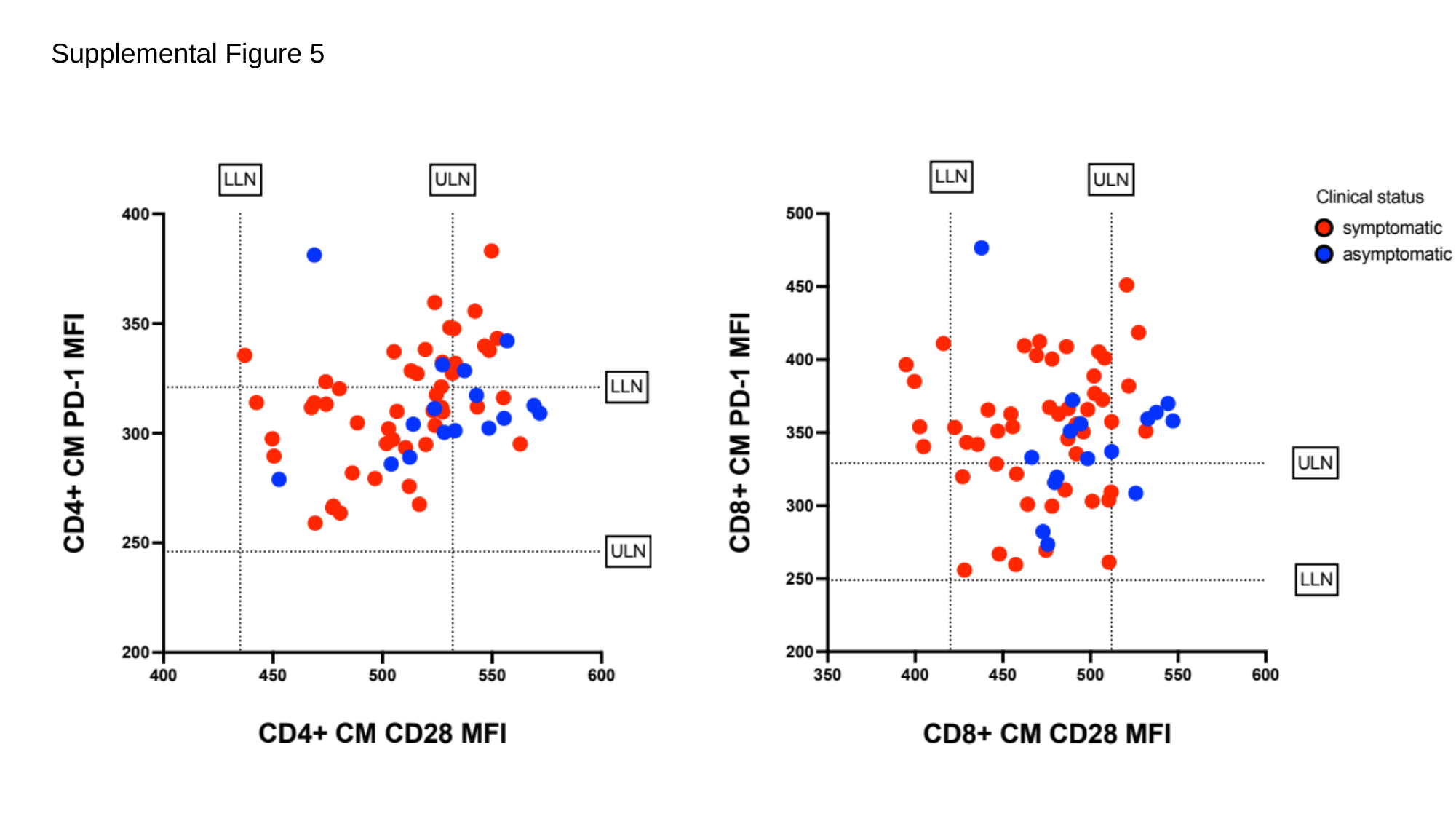

Supplemental Figure 5
